# Unravelling the epidemiological and dispersal dynamics of the 2024-2025 chikungunya virus epidemic on Réunion island

**DOI:** 10.64898/2026.01.07.26343606

**Authors:** Etienne Frumence, Raphaëlle Klitting, Kyla Serres, Yucai Shao, Filippo Monti, Muriel Vincent, Mandev S. Gill, Marc A. Suchard, Philippe Lemey, Xavier de Lamballerie, Marie-Christine Jaffar-Bandjee, Simon Dellicour

## Abstract

Réunion island experienced a massive chikungunya virus epidemic in 2024-2025, with >54,000 confirmed cases. This is the second major chikungunya epidemic on the island, following the first one that peaked 20 years ago. It has been asserted that this new outbreak finds its origin in a single introduction event into the island, offering an opportunity to exploit viral genomic data to understand the epidemiological and dispersal dynamics of the introduced transmission chain. We sequenced >3,000 viral genomes collected during the epidemic. Harnessing this genomic dataset, we used several phylogeographic and phylodynamic approaches to unravel the paths taken by the transmission chain and the external factors that might have impacted its dispersal and epidemiological dynamics on the island. Our analyses highlight a dispersal pattern in line with a gravity-model dynamic with viral transition events being more frequent from and toward more populated areas. Our analyses also reveal that the transmission chain was overall spatially intermixed, with frequent exchanges among residential areas. In addition, we show that the temporal dynamic and intensity of the epidemic were associated with climatic variables, namely temperature and precipitation. Our results also show that, in theory, the population immunity – resulting from this epidemic and the previous one (2005-2006) – could be sufficient to explain on its own the decrease in the transmission rate that led to the end of the epidemic. While a short-term resurgence cannot be excluded, the risk of a large-scale circulation of the virus in the human population appears therefore relatively limited in the upcoming seasons.

---

Chikungunya virus (CHIKV) is a positive-sense, single-stranded RNA *Alphavirus* of the Togaviridae family transmitted to humans by the bite of infected *Aedes* mosquitoes. CHIKV is divided into three main phylogenetic genotypes: the West African lineage, currently limited to West Africa, the Asian lineage, which circulates in both Asia and the Americas, and the East/Central/South African (ECSA) lineage, which is the most widely distributed taxon (1). Historically, the virus circulated in African enzootic cycles involving non-human primates, but it can also spread in urban and peri-urban environments due to the anthropophilic nature of *Aedes aegypti* and *Aedes albopictus* (2). In the current globalised context, CHIKV has emerged as a major international public health concern – CHIKV outbreaks having been detected in over 100 countries across tropical and sub-tropical areas since the 2000s (3) – with human mobility, together with the widespread expansion of competent vectors, playing a crucial role in its spatial expansion (4, 5). Millions of cases were reported globally over the last decade (6). While associated with a low fatality rate (<1/1000), CHIKV infection causes substantial morbidity with ∼85% of patients exhibiting high fever (>39°C), severe arthralgia/myalgia, and an erythematous, maculopapular rash (1). Furthermore, a substantial proportion of patients develop long-term musculoskeletal symptoms (1), including persistent and often disabling joint pain lasting months to years – after the first Réunion epidemic, up to 60% of patients had relapsing arthralgia up to 36 months post-infection (7, 8).

Between August 2024 and June 2025, Réunion island experienced a notable CHIKV epidemic, with >54,000 confirmed cases and 43 deaths (*Santé Publique France - la Réunion* epidemiological updates (9, 10)), making it the second major CHIKV epidemic recorded on the island. The first CHIKV episode on Réunion occurred in 2005-2006 and affected an estimated ∼34% of the population (11, 12), with occasional circulation detected up to 2011 (13, 14). This second major epidemic stems from a new introduction, after more than ten years with no autochthonous cases reported on the island (15). As this new epidemic likely finds its origin in a single introduction event into the island (15), it offers a unique opportunity to exploit viral genomic data to understand the epidemiological and dispersal dynamics of the introduced transmission chain. During the first half of 2025, the 2024-2025 Réunion epidemic lineage further spread to other Indian Ocean islands, including Mayotte and Mauritius (15), and, in the summer of 2025, led to unprecedented numbers of cases in more temperate regions of Europe (16) (France ∼780 cases, Italy ∼380 cases) and in China (17) (>16,000 cases in Guangdong province).

Throughout the epidemic, in-depth genomic surveillance was ensured by the associated Arbovirus French National Reference Center (NRC) in Réunion, with (i) systematic sequencing of all CHIKV-positive cases transferred to the NRC from August 2024 to mid-February 2025, and (ii) sequencing of random subsamples after mid-February, due to the sharp increase in case numbers. This considerable effort allowed the generation of more than 3,000 near-full viral genomes sampled over the entire epidemic across the island, representing over 5% of confirmed cases, a genomic sampling fraction only achieved for a few other pathogens, including Ebola (18) (∼5%) and SARS-CoV-2 (reaching above 10% in several countries, including the United Kingdom and Denmark (19)). This large-scale dataset provides an exceptional basis to track the evolution, population dynamics and spatial spread of viral lineages during the epidemic.

Here, we leverage this comprehensive genomic dataset to understand the epidemiological dynamics of the 2024-2025 CHIKV epidemic on Réunion island. Our first aim was to investigate the main factors associated with the temporal dynamic and the end of the epidemic. For this purpose, we employed phylodynamic approaches to test for statistical associations with climatic factors and to investigate the contribution of the level of population immunity reached across the island to the decrease in transmission rate having led to the end of the epidemic. While climatic conditions (temperature, precipitation) seem to be associated with the evolution of the size of the viral population, we also highlight that population immunity could theoretically be sufficient to explain on its own the effective reproduction number falling below one at the end of the epidemic. This result suggests that there is a limited risk of epidemiological rebound when climatic conditions become suitable again in the upcoming seasons. The second aim of our study was to unravel the dispersal dynamics and drivers of the viral transmission chain once introduced on the island. Our phylogeographic approach reveals a spatially intermixed transmission chain with frequent viral exchanges between the different municipalities, with more populated residential areas preferentially attracting and seeding dispersal events to other locations. From a public health perspective, these results provide actionable insights for anticipating future CHIKV epidemics.

## Results

To investigate the epidemiological and dispersal dynamics of CHIKV during the 2024-2025 epidemic, we sequenced a total of 3,114 near-full viral genomes sampled between August 2024 and August 2025, all geo-referenced and associated with a precise collection date (Figs. 1A-B, S1). The resulting sequence alignment constitutes a comprehensive genomic dataset representative of the epidemic, with a number of CHIKV genomes collected per week highly correlated with the weekly number of confirmed cases (Pearson coefficient = 0.95; Fig. S2A), and also showing a good correlation between the total numbers of collected genomes and reported cases per municipality (Pearson coefficient = 0.86; Fig. S2B). Our preliminary phylogenetic analysis – based on a restricted set of Réunion sequences and a selection of international sequences – confirms the monophyletic nature of the Réunion clade, in line with the hypothesis of an epidemic initiated by a single introduction event into the island (Fig. S3). As identified previously, this clade belongs to CHIKV ECSA-2 genotype and arises from an African clade (Fig. S3), which mostly gathers sequences from the Central African region (15).

**Figure 1.**
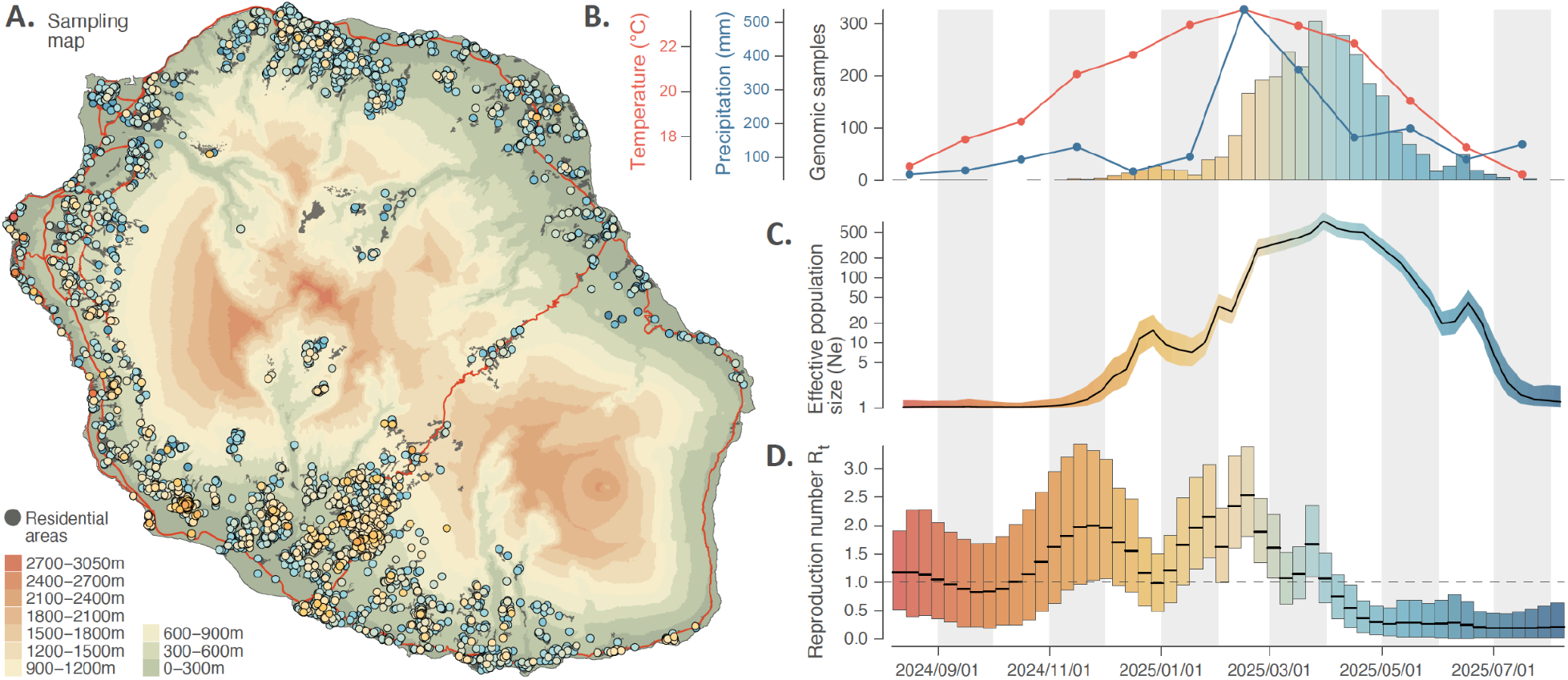
Sampling map and phylodynamic analyses of the 2024-2025 chikungunya virus (CHIKV) epidemic on Réunion island. **A**: topographic map of the island displaying the spatio-temporal distribution of the CHIKV genomic samples collected and sequenced during the epidemic (dots coloured according to sampling date). On this map, red lines and grey areas correspond to the main roads and residential areas, respectively. **B**: distribution of the number of sampled CHIKV genomes on a weekly basis (histogram coloured according to time), as well as the evolution of monthly average of the daily mean temperature (red curve) and monthly cumulative precipitation (blue curve) during the course of the epidemic. **C**: dynamics through time of the overall effective viral population size (Ne) as estimated with a phylodynamic analysis based on sampling-aware skygrid analysis; the grey curve and surrounding ribbon coloured according to time correspond to the posterior median estimate and associated 95% highest posterior density (HPD) intervals, respectively. **D**: dynamics through time of the effective reproduction number (R_t_) as estimated with a phylodynamic analysis based on an episodic birth-death-sampling model; with vertical boxes coloured according to time corresponding to weekly 95% HPD intervals, and the grey thick lines to weekly posterior median estimates (see the text for further detail on both the sampling-aware skygrid and episodic birth-death-sampling analyses).

We then conducted a series of time-scaled phylodynamic and phylogeographic analyses based on the full CHIKV genomic alignment assembled for the Réunion epidemic to understand the epidemiological and dispersal dynamics of the virus and associated drivers (see the Methods section for further detail on these approaches and models, as well as Figure S4 for a visualisation of a time-scaled phylogenetic inference). First, we performed a time-scaled phylogenetic inference using a sampling-aware skygrid coalescent model (20) to estimate the evolution of the effective viral population size (Ne) during the epidemic. The effective population size represents the size of an idealised population (e.g., with no spatial structure) whose trajectory over time can serve as a proxy for changes of the actual viral population. This analysis reveals a bell-shaped curve (Fig. 1C) in phase with the epidemiological curve, coinciding with the end of the epidemic around July 2025.

To investigate how temporal variations in effective viral population size could be explained by changes in transmission intensity during the epidemic, we estimated the weekly evolution of the effective reproduction number (R_t_; the average number of persons infected by a positive case at a specific point in time). Still based on our CHIKV genomic data, we conducted a time-scaled phylogenetic inference using an episodic birth-death sampling model (21). This analysis reveals two main epidemic phases during which R_t_ remained consistently above one for several weeks in a row (Fig. 1D): from mid-October to mid-December 2024 and from mid-January to early March 2025; these two phases correspond to periods of increasing effective viral population size. These weekly estimates also clearly indicate that from April 2025 onward, R_t_ drops and stays below one, indicating the decline of the epidemic with one infected person generating on average less than one secondary case (22).

The temporal variations in Ne and R_t_ captured in the above analyses show epidemiological trends that appear associated with the temporal variation in climatic covariates (Fig. 1B-D). Regarding the effective population size, the peak of the curve follows by 1-2 months the peaks in temperature and precipitation that correspond to the height of the rainy season. Similarly, the estimated drop in R_t_ appears to align with the decrease in temperature and precipitation that marks the end of the rainy season. This temporal association raises the question of the contribution of the decline in temperature and precipitation to the subsidence of the CHIKV epidemic, especially as these factors influence the activity and density of mosquito vectors (23–27). A similar observation can be made for the two epidemic waves of the previous 2005-2006 CHIKV epidemic on the island, which occurred in March-June (2005) and December-April (2006), respectively (28). These time frames roughly correspond to the same period of the year, both broadly overlapping with the rainy season, which further suggests that variations in temperature and precipitation could be associated with the temporal dynamic of CHIKV epidemics on the island (as formally tested below). Finally, while such climatic factors could influence arbovirus transmission dynamics, the level of population immunity reached on the island could also be another key variable shaping the virus transmission dynamic and intensity.

To formally test for associations between the epidemiological dynamic of the epidemic and putative explanatory covariates, we used two phylodynamic approaches. However, as seroprevalence estimates were not available through time, we focused our first analyses on climatic covariates. Specifically, we tested the associations between two climatic variables – monthly average of daily mean temperatures and monthly cumulative precipitation – and both monthly viral effective population size (Ne) and effective reproduction number (R_t_). To this end, we used multivariate generalised linear model (GLM) extensions of the sampling-aware skygrid coalescent model (hereafter referred to as “skygrid-GLM” analyses) and of the episodic birth-death-sampling model (referred to as “EBDS-GLM” analyses), respectively (29). The skygrid-GLM analyses reveal a statistically supported positive association between effective population size and both covariates (Fig. 2A-B), with 95% highest posterior density (HPD) intervals of GLM effect size coefficients that consist entirely of positive values. To investigate the possibility of a delayed association between temporal changes in the effective population size with covariate fluctuations, we also conducted this analysis while considering the association between the monthly effective population size and the covariate values corresponding to the preceding month. The results once again reveal statistically supported positive associations with both climatic covariates (Fig. 2C-D). Of note, the small variations in the effective population size around December 2024 and June 2025 (Fig. 1C) could be related to fluctuations of a climatic factor such as precipitation. With the EBDS-GLM analyses, only the 95% HPD interval associated with the GLM coefficient of the temperature covariate excludes zero, confirming a statistically supported positive association between the temperature and R_t_ (Fig. 2E-F). This supported association is however not identified when considering a one-month lag period (Fig. 2G-H). These results confirm that during the 2024-2025 epidemic, temperature and precipitation were correlated with the evolution of the effective population size Ne and, solely in the case of temperature, with the effective reproduction number R_t_, suggesting a potential causal link between these climatic variables and virus transmission dynamics.

**Figure 2.**
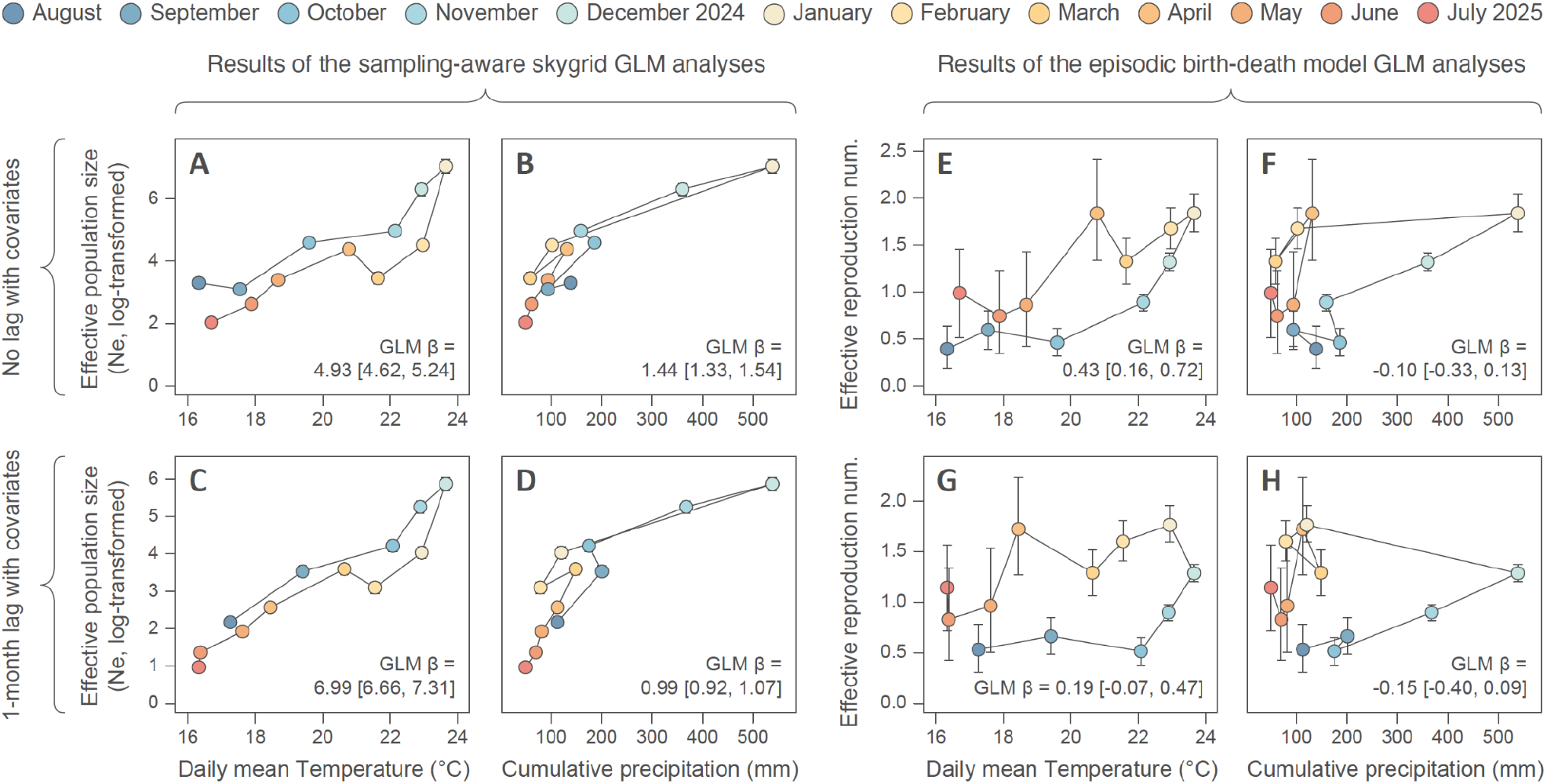
Analyses of the associations between climatic covariates and the epidemiological dynamic of the 2024-2025 chikungunya virus (CHIKV) epidemic on Réunion island. Specifically, we tested the associations between two climatic variables – monthly average of daily mean temperatures or monthly cumulative precipitation – and both monthly viral effective population size (Ne) and the effective reproduction number (R_t_). To this end, we used generalised linear model (GLM) extensions of the sampling-aware skygrid coalescent model (referred to as the “skygrid-GLM” analyses) and of the episodic birth-death-sampling model (referred to as the “EBDS-GLM” analyses), respectively. We here display the relationship between monthly climatic values and monthly Ne (obtained from the joint analysis of genomic sequence and climatic covariate data; **A-D**) or monthly R_t_ (**E-H**); Ne and R_t_ posterior median estimates being displayed along their 95% highest posterior density (HPD) interval (which is not always visible when too small). In addition, we also report the posterior median and 95% HPD interval of the GLM coefficient (β) associated with each climatic covariate and analysis. While the first row of graphs report the results of the analyses conducted without considering a time lag, the second row of graphs report the analyses based on a one-month lag period between the monthly climatic covariates and Ne or R_t_ (each monthly Ne and R_t_ being then associated with the climatic value of the previous month). A statistically supported association between the covariate and Ne or R_t_ is inferred when the 95% HPD interval of the GLM coefficient excludes zero.

Due to an absence of time-resolved seroprevalence data, we could not assess the association between the evolution of population immunity and epidemiological parameters (Ne, R_t_). However, this variable is likely also contributing to transmission dynamics and constitutes a key element to determine the risk of epidemiological rebound when climatic conditions become suitable again during the next rainy season. We thus conducted a theoretical exercise to investigate whether the R_t_ drop revealed by our episodic birth-death model analysis could also be related to the island population reaching sufficient herd immunity. To address this question, we started by estimating the exponential growth rate of the epidemic with a third time-scaled phylogenetic inference, this time using an exponential growth coalescent model. This exponential growth rate estimate was subsequently used to calculate the epidemic basic reproduction number (R_0_; the average number of persons infected by a positive case at the beginning of the epidemic) (30). With this approach, we estimate that the R_0_ value ranges from 1.39 (95% HPD = [1.36, 1.41]) to 2.27 (95% HPD = [2.18, 2.38]), which is in line with our R_t_ estimates during the early stages of the epidemic. Yet, this range of R_0_ values is lower than estimates reported by some (28, 31, 32), but not all (33, 34) studies using epidemiological modelling approaches to estimate R_0_ for the previous Réunion epidemic of 2005-2006 (35). We then exploited this estimated range of R_0_ values together with seroprevalence data to estimate the effective reproduction number at the end of the epidemic using the following formula (22): R_t_ = (1 −p_C_) (1 −p_I_) R_0_, where p_C_ and p_I_ are the relative reduction in transmission rates due to non-pharmaceutical interventions and the proportion of immune individuals, respectively (22). Still in the context of a theoretical exercise, we estimate R_t_ under the simplified assumption of no transmission rate reduction due to intervention strategies (p_C_ = 0), and we estimate p_I_ using seroprevalence data published in November 2025 by *Santé Publique France* (36). According to this report, the global seroprevalence at the scale of the island is estimated to be 66.0%, which would result from residual immunity following the 2005-2006 epidemic (∼20%) and from the immunity acquired during the 2024-2025 epidemic (∼46%) (36). Assuming that the seroprevalence estimate corresponds to the proportion of immunised people, we estimate a pI value of 0.575 obtained by the ratio between (i) the proportion of new people immunised after the 2024-2025 epidemic (∼46%) and (ii) the proportion of the population non-immunised at the beginning of it (∼80%). Using the estimated range of R_0_ values, we can then estimate an effective reproduction number at the end of the epidemic ranging from 0.59 (95% HPD = [0.58, 0.60]) to 0.97 (95% HPD = [0.93, 1.01]). This result confirms that the current population immunity, as estimated by prevalence data, could in theory be sufficient to explain the R_t_ dropping below one and the end of the epidemic.

Our first set of analyses identifies associations between epidemiological and climatic trends as well as population immunity as likely key contributors to the end of the 2024-2025 Réunion epidemic, but it does not investigate the factors that drive the spatial spread of the virus during the epidemic. To address this question, we capitalise on our genomic dataset with a spatio-temporal distribution that is representative of the overall epidemic (Fig S2A-B) to conduct a fine-scale phylogeographic reconstruction of the virus dispersal history between municipalities (Fig. 3). The first chikungunya cases were detected mid-2024 in the western part of the island, in the municipality of Saint-Paul. By the end of 2024, the virus spread to southwestern municipalities (Saint-Pierre, Le Tampon; Fig. 3). In early 2025, the virus subsequently spread towards most urban centers across the island, while maintaining a more intense circulation in the southwestern region. During the course of the epidemic, southwestern municipalities — in particular Le Tampon — have also remained an important source of viral lineage dispersal events towards other municipalities across the island. As illustrated during the peak of the epidemic around March and April 2025, frequent back-and-forth exchanges of viral lineages occurred between several pairs of municipalities, highlighting the important interconnectivity of residential areas within the transmission chain (Fig. 3). Overall, our analysis reveals that the transmission chain is notably intermixed between the different municipalities. This is further highlighted by the relatively high normalised Shannon entropy measures estimated for each municipality. Ranging from 0 (corresponding to a single municipality cluster) to 1 (each sample in the municipality corresponds to a distinct introduction event in that municipality), this metric can be used to indicate how phylogenetically structured the epidemic was in each location (37). Here we estimate and get statistical support for a normalised entropy higher than 0.5 for 19 (∼80%) and higher than 0.75 for 13 (∼54%) out of 24 municipalities (Fig. S5). Interestingly, the two municipalities associated with the lowest entropy values (L’Étang Salé and Le Tampon, with a posterior median estimate <0.5) are the two first municipalities strongly affected by the epidemic (Fig. 3). We can also note that some municipalities considered as less urbanised and/or geographically more isolated – such as Cilaos, Petite-Île, Saint-Joseph, and Saint-Philippe – are also associated with a relatively lower entropy value (posterior median estimate <0.75; Fig. S5).

**Figure 3.**
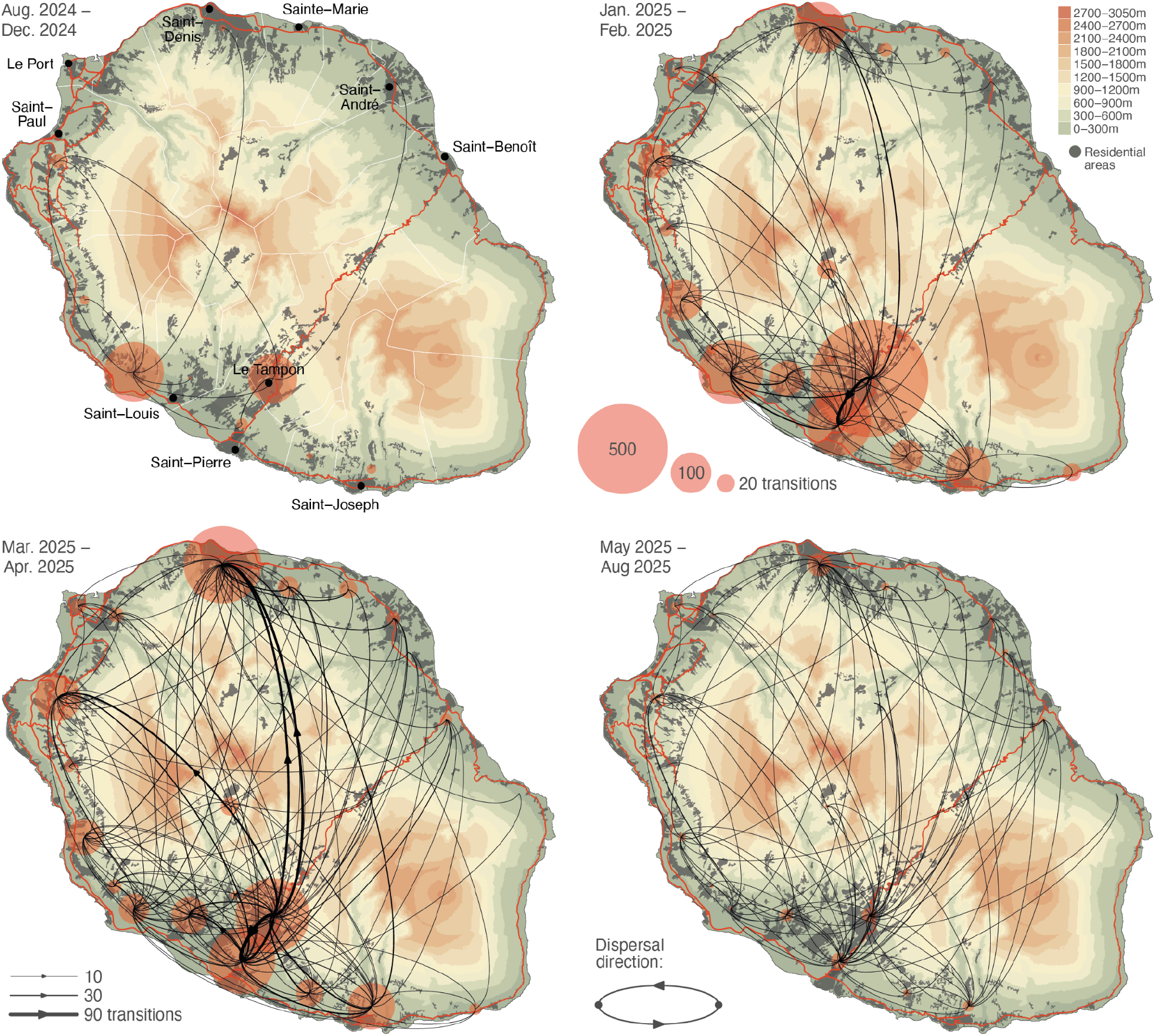
Phylogeographic analysis of the dispersal history of viral lineages during the 2024-2025 chikungunya virus (CHIKV) epidemic on Réunion island. The maps correspond to four successive snapshots displaying the discrete phylogeographic reconstruction of the dispersal history of CHIKV lineages conducted at the municipality level. On these maps, municipalities are associated with a transparent red dot with the size being proportional to the number of inferred lineages within the considered municipality (each dot having been placed at the centroid point computed from the sampling locations of all genomic sequences collected in the municipality), and curved arrows illustrate with their thickness the expected number of viral lineage transition events inferred from one municipality to another during the considered time period. These topographic maps are coloured according to the altitude or in grey when corresponding to residential areas. Red lines correspond to the main roads on the island and, solely reported on the first map, white lines correspond to the borders of the municipalities. On this first map, we also report the position and names of the main urban areas (dark grey dots). See also the GitHub repository associated with the present study for an animation of the discrete phylogeographic reconstruction generated with the spread.gl program (38).

To identify the factors driving the viral spread and circulation across the island, we conducted another multivariate analysis, this time coupling phylogeographic analysis with a GLM aiming at evaluating the potential predictors of viral spread between sampled locations. With Bayes factor values higher than 20, we find strong support for (i) a negative association between geographic distance and lineage transition frequency among locations (GLM coefficient = -0.48, with a 95% HPD interval = [-0.56, -0.32]), and for (ii) positive associations with the population size at both the location of origin (GLM coefficient = 0.51 [0.33, 0.63]) and destination (GLM coefficient = 0.98 [0.81, 1.10]) of lineage transition events (Table 1). This analysis reveals that viral lineage transition events occurred more frequently between municipalities that are geographically closer, as well as from and towards municipalities with a higher population count. Together, these results highlight a gravity-model dynamic, where dispersal between locations is a function of their population sizes and the spatial distances between them (18). We further assessed the impact of geographic distance by estimating the isolation-by-distance (IBD) signal associated with the viral spread. Here computed as the correlation between the evolutionary and geographic distance (39), the IBD signal can be used to measure to what extent the geographic distance can explain the divergence between sequenced cases within the transmission chain. With a mean Pearson correlation of 0.11 (95% HPD = [0.10, 0.13]), this analysis reveals a significant yet relatively low IBD signal. This result indicates that while geographic distance had a significant impact favouring higher dispersal frequencies of viral lineages among nearby locations, it was at the same time however not a strong determinant of the risk of viral spread among locations. Based on these results, human mobility could arise as a potential predictor of choice to explain why specific pairs of municipalities had more frequent viral lineage exchanges than others. However, with a limited effect size and a Bayes factor equal to 11, we only find a positive yet not strong statistical support (40) for an association between the frequency of viral transition events and the professional commuting flow estimated between municipalities (Table 1). While other non-tested predictors could contribute, it is also possible that the professional mobility data used here to estimate the human flow do not fully capture the mobility spectrum between municipalities.

**Table 1.** Multivariate analysis of the predictors of the transition frequencies of viral lineages among municipalities during the 2024-2025 chikungunya virus (CHIKV) epidemic on Réunion island. We report the results of the generalised linear model (GLM) multivariate analysis coupled with a discrete phylogeographic analysis to jointly infer the transition events between municipalities and the association between transition frequencies and several potential predictors; namely four covariates: the geographic distance and professional commuting flow between municipalities, as well as the human population count at the municipality of origin and destination. Support for each predictor is computed and reported by its inclusion probability, i.e. the frequency across an MCMC chain at which the predictor is included in the model (E[δ]), which can also be translated into a Bayes factor (BF) support. Finally, the contribution of each predictor to the model is reported by the posterior mean and 95% highest posterior density (HPD) interval of its GLM coefficient (β) conditional on the predictor being included in the model (β|δ= 1).

| Covariate | Inclusion probability ($E[\delta]$) | Bayes factor (BF) | GLM coefficient $\beta \mid \delta=1$ |
| --- | --- | --- | --- |
| Geographic distance | 1.000 | >99 | -0.476 [-0.562, -0.320] |
| Professional commuting flow | 0.500 | 11.0 | 0.029 [0.000, 0.478] |
| Population at origin | 1.000 | >99 | 0.514 [0.331, 0.630] |
| Population at destination | 1.000 | >99 | 0.980 [0.813, 1.096] |

## Discussion

Genomic surveillance during the 2024-2025 CHIKV epidemic on Réunion island has led to the generation of >3,000 geo-referenced viral genomes with detailed time-stamps, constituting one of the most comprehensive genomic datasets for a single epidemic (excluding SARS-CoV-2). When coupled with detailed metadata of sampling date and location, genomic samples collected throughout the course of an epidemic allow us to infer evolutionary connections among cases, and as a result, to unravel the paths and dynamics of the transmission chain, leading to a clear picture of the past and current epidemiological patterns. In this study, we harness this comprehensive dataset of CHIKV genomes from Réunion 2024-2025 to conduct a series of state-of-the art phylodynamic and phylogeographic investigations. From an analytical perspective, we highlight the value of such a comprehensive genomic dataset to provide detailed insights into the viral epidemic and dispersal dynamics, as well as the factors driving these dynamics. Investigating these drivers is of particular interest for arthropod-borne viruses such as chikungunya, as virus transmission can be affected by factors related to the human host but also those influencing their invertebrate vector.

Our analyses confirm an evolutionary pattern coherent with a main single introduction event at the origin of the 2024-2025 chikungunya epidemic on Réunion island. While we cannot exclude subsequent re-introductions from contaminated neighbour islands/countries, our results confirm that the transmission chain circulating across the island originated from a single ancestor that traces back to the central African lineage. Once introduced, the circulation of the virus within the island has followed a gravity-model dynamic, with more frequent dispersal events from and towards more populated areas as well as between geographically close areas. Such a gravity-model dynamic was for instance previously identified in the context of the 2013-2016 Ebola virus epidemic that occurred in West Africa (18). In the meantime, our analyses also reveal a limited association between the spatial distance and the divergence between two cases within the transmission chain. These results are however not necessarily in contradiction: while the discrete phylogeographic analysis coupled with a GLM reveals that smaller geographic distances are significantly associated with more frequent dispersal events, the isolation-by-distance signal analysis however shows that the distance between locations is not a strong determinant of the risk of viral transition events among locations. Overall, our results highlight an important intermixing of the transmission chain between the different municipalities. This is illustrated in particular at the peak of the epidemic, with frequent dispersal events between distinct residential areas, likely sustained by an important human – and thus viral – mobility within the island. Differences in vector competence across the island, as has been reported for dengue (41), could also have influenced virus dispersal by favouring transmission in certain municipalities over others. However, in the absence of reliable competence data for CHIKV at the municipal level, we were unable to formally evaluate this impact.

Finally, our analyses also reveal that the progressive buildup of herd immunity within the human population could in theory have been sufficient to explain the decline in the effective reproduction number below one. Other factors might however also have contributed to bringing the epidemic to an end. In particular, and as also formally tested by our phylodynamic analyses, estimated variations in the effective viral population size and reproduction number are positively associated with the evolution of temperature, and also precipitation in the case of the effective population size; two variables that have been shown to affect vector density, activity and vectorial transmission (23– 27). Of note, a vaccination campaign with the IXCHIQ® (VLA1553, Valneva SE) was launched in April 2025 on the island, but it was soon hindered by the identification of severe adverse events following vaccination (42), which led to a temporary suspension of vaccination in people over 65 years by the French *Haute Autorité de Santé* and to a full suspension of the vaccine by the US Food and Drug Administration (FDA) in August 2025. As only about 10,000 vaccine doses were administered on the island in 2025 (data from the Regional Health Agency – *ARS La Réunion*), and as those were mostly used after the peak of the epidemic, we expect a minimal contribution of IXCHIQ vaccination to curbing virus transmission. Overall, with the notable population immunity (66%) apparently reached by the Réunion population, the risk of severe resurgence of viral circulation appears to be low, and, in fact, was not observed during the following season, with only 46 local cases reported as of June 2026 (source: www.santepubliquefrance.fr), in spite of the substantial CHIKV circulation reported on other islands of the Indian Ocean, including Mauritius and the Seychelles (source: www.who.int).

In conclusion, we achieve an in-depth investigation of the epidemiological and dispersal dynamics of the second main CHIKV epidemic on Réunion island applying phylodynamic and phylogeographic approaches to an extensive dataset of virus genomes. We identify several key human-dependent and climatic factors that shaped the epidemic, with human mobility and density contributing to virus spread, while the end of the rainy season and the progression of population immunity likely contributed to curb virus transmission. Our results provide a first detailed picture of this epidemic, guiding vector-control measures and immunisation strategies on the island for future events of local transmission of CHIKV or other vector-borne diseases including dengue virus. This first set of analyses can be combined with further investigations of vector-dependent factors (including vector density and competence) to achieve a systemic comprehension of the factors that influence virus transmission.

## Materials and Methods

### Ethical statement

Samples included in this study consisted of residual serum or plasma from chikungunya cases referred to the NRC for arboviruses in Réunion island. Sequencing data were generated as part of routine surveillance activities within the national public health surveillance program coordinated by the NRC and supervised by the National Public Health Agency (*Santé Publique France*). As an epidemiological record, consultation with an ethics committee was not required. No additional clinical specimen was collected for the purpose of this study. Human samples were anonymised, with no or minimal risk to patients, in accordance with the European General Data Protection Regulation (GDPR) and the French National Commission on Informatics and Liberty (CNIL).

### Sampling and genomic sequencing

CHIKV-positive clinical samples were analysed as part of the island-wide arbovirus genomic surveillance program coordinated by the associated Arbovirus French National Reference Center (NRC) in Réunion (15). Located at the northern site of Réunion Island University Hospital, the NRC automatically receives all arbovirus-positive samples from both the northern and southern hospital sites; the other public and private laboratories of the island refer their samples to the center. From August 2024 to mid-February 2025, all confirmed CHIKV-positive samples detected by public and private laboratories were forwarded to the NRC for complementary analyses whenever available. Thereafter, due to increasing case numbers, a randomised weekly subset (up to 30 samples) was selected under the coordination of health authorities (*Santé Publique France* at Réunion island) to ensure a geographically representative sampling for each laboratory group. Samples processed at Réunion Island University Hospital were systematically included throughout. Part of the samples from the Western Hospital Center, which covers roughly one quarter of the island’s population, could not be retrieved during the epidemic peak (fewer than 200 positive samples) and were therefore not included in this study. Whole protein-coding sequences were generated using an in-house amplicon-based protocol on the MinION sequencing platform (Oxford Nanopore Technologies), including RNA extraction from serum or plasma, two-pool multiplex RT-PCRs using a custom CHIKV primer scheme, and barcoding with the Rapid Barcoding Kit V14, as previously described (15). From April 2025 onward, an updated primer scheme incorporating degenerated oligonucleotides was adopted to improve amplicon balance and to rescue weak or failed regions, particularly amplicon 2. The complete modified primer list is provided in Table S1. Multiplex PCRs using the updated primers were performed under the following cycling conditions: 98°C for 30 s, then 40 cycles of 98°C for 15 s, 58°C for 2 min, and 72°C for 3 min, followed by a final extension at 72°C for 3 min. Sequencing was performed on R10.4.1 flow cells, and consensus genomes were generated using the ARTIC field bioinformatic pipeline (v1.6.1). Only assemblies with more than 80% of the coding region covered at a minimum depth of 100x and originating from patients residing on Réunion Island with a confirmed residential address were included in this study. In total, we sequenced 3,114 CHIKV genomes sampled from the 2024-2025 Réunion epidemic (Figs. 1, S1), which were deposited on GenBank with accession numbers PV035814, PV685534-PV685706, and PX797910-PX800869.

### Preliminary phylogenetic analyses

We first downloaded all publicly available CHIKV sequences from the GenBank database (keywords: “chikungunya virus”; accessed on April 19, 2025) and then filtered the data by discarding (i) sequences from laboratory strains (adapted, passaged multiple times, obtained from antiviral or vaccine experiments), (ii) sequences that did not belong to the CHIKV species, and (iii) sequences covering less than 85% of the viral genome. The remaining 2,150 sequences were combined with our 3,114 sequences from the 2024-2025 Réunion epidemic, aligned using MAFFT v7.511 (43), inspected manually using the program AliView v1.0 (44), and trimmed to their ORFs. We then performed a maximum-likelihood (ML) phylogenetic reconstruction with IQ-TREE v1.6.12 (45) using the best-fit substitution model identified by ModelFinder (46) and assessed branch support using an ultrafast bootstrap approximation (UFBoot2; with 1,000 replicates). Based on this first ML phylogenetic reconstruction, we selected the 200 sequences most closely related to the 2024-2025 Réunion epidemic sequences to constitute an evolutionary background for subsequent phylogenetic analyses. These 200 background sequences were combined with 53 sequences subsampled from the full set of CHIKV genomic sequences generated from the 2024-2025 Réunion epidemic by randomly selecting five sequences per month. With this resulting set of 253 sequences, we performed another phylogenetic reconstruction as described above, followed by a root-to-tip regression analysis conducted with the program TempEst (47) to assess the temporal signal associated with the genomic dataset (R^2^ = 0.67; Fig. S2C). Following the root-to-tip regression analysis, we discarded two background sequences whose sampling date was likely incongruent with their genetic divergence, resulting in a subset of 251 sequences exhibiting sufficient association between genetic distances and sampling dates (R^2^ = 0.71; Fig. S2D).

### Time-scaled phylogenetic inferences

The resulting alignment of 251 sequences — 53 sequences from the Réunion epidemic and 198 background sequences (see above) — was used to conduct a first time-scaled phylogenetic inference aiming (i) to assess the monophyletic nature of the Réunion clade and (ii) to estimate a substitution rate for CHIKV (Fig. S3A). All time-scaled phylogenetic inferences (including the following phylodynamic and phylogeographic inferences) were conducted with the software package BEAST X v1.10.5 (48) using a GTR+Γ (general time-reversible with discretised gamma-distributed rate heterogeneity) nucleotide substitution model (49). For this first time-scaled phylogenetic analysis, we set a relaxed molecular clock with an underlying log-normal distribution to model branch-specific evolutionary rates (50) and a flexible skygrid coalescent model for the tree prior (51). We ran the analysis for 6 x 10^8^ Markov chain Monte Carlo (MCMC) iterations, sub-sampling posterior trees and parameter values every 100,000 iterations, and eventually discarding the initial 10% of sub-samples as burn-in. We assessed the MCMC convergence and mixing with the program Tracer v1.7 (52), checking that all continuous posterior estimates were associated with an effective sample size value >200. The maximum clade credibility (MCC) tree was retrieved and annotated with the program TreeAnnotator v1.10.5 (48), and plotted in R with a custom script (see the *Code and data availability* section).

With this first analysis, we estimated a mean posterior substitution rate of 4.095 x 10^-4^ substitutions/site/year (s/s/y; 95% highest posterior density [HPD] interval = [3.312 x 10^-4^, 4.860 x 10^-4^]), which broadly aligns with previous estimates of CHIKV mean evolutionary rate ranging from ∼2 to ∼8 x 10^-4^ s/s/y (15, 53, 54). The resulting posterior distribution (Fig. S3B) was used to inform a strict molecular clock model for all subsequent time-scaled phylodynamic and phylogeographic analyses performed on the complete CHIKV genomic alignment for the Réunion epidemic. Specifically, for these analyses, we set an informative prior for the substitution rate parameter, using a normal distribution defined by the mean and standard deviation values retrieved from the posterior distribution inferred by the first time-scaled phylogenetic inference based on a restricted but international genomic dataset; i.e. a mean and a standard deviation of 4.095 x 10^-4^ and 4.039 x 10^-5^ s/s/y, respectively. Without such an informative substitution rate prior, time-scaled phylogenetic inferences based on the Réunion epidemic alignment would fail to converge (data not shown); likely due to the relatively limited genetic variability associated with the Réunion epidemic alignment, which makes calibrating a molecular clock model solely on this alignment challenging.

Taking advantage of this informative substitution rate prior, the second time-scaled phylogenetic inference was based on the overall CHIKV alignment for the Réunion epidemic (*n* = 3,114 genomic sequences) and aimed (i) to generate a posterior distribution of time-scaled phylogenetic trees to be used as an empirical distribution of tree topologies for the phylogeographic inference (see below) and (ii) to estimate the exponential growth rate of the epidemic. For the second time-scaled phylogenetic analysis, we set an exponential growth coalescent model for the tree prior in BEAST X and ran the analysis for 7 x 10^8^ MCMC iterations while sub-sampling posterior trees and parameter values every 100,000 iterations. Upon discarding the first 10% of the sub-samples as burn-in, we assessed the MCMC convergence and mixing, as well as retrieved, annotated, and visualised the MCC tree as described above. The exponential growth rate (r_0_) estimated by this analysis was in turn exploited to estimate the basic reproduction number (R_0_) associated with the epidemic (30, 55). For this purpose, we assumed a gamma-distributed serial interval (i.e. the time between the symptoms onset of a case and that of the related secondary cases) and implemented an iterative procedure whereby multiple plausible serial interval (SI) distributions were sampled and used to re-estimate R_0_, which allowed propagating the uncertainty associated with the SI of CHIKV. Specifically, for each r_0_ value sampled from the posterior distribution, we conducted 1,000 iterations for which the SI mean was drawn from a uniform distribution ranging from 9 to 23 days and the SI standard deviation from a uniform distribution ranging from 4 to 8 days, reflecting a range of values reported and considered in the literature (56–59).

### Inferring the effective viral population size

We inferred the dynamics of the effective size of the overall viral population during the Réunion epidemic (Fig. 1C) using a sampling-aware generalisation of the skygrid coalescent model (20) (hereafter referred to as the “sampling-aware skygrid” model) implemented in the software package BEAST X v1.10.5 (48). Standard coalescent-based models implicitly assume that there is no dependence between the effective population size and the distribution of sequence sampling times. However, it is possible that, for instance, viral samples are collected more frequently when the effective viral population size is high, and failure to account for such preferential sampling can lead to biased inference of population dynamics. The sampling-aware skygrid model overcomes such difficulties by modelling sequence sampling times as a Poisson process whose intensity is a log-linear function of the log effective population size (20). For the sampling-aware skygrid analysis, we ran and eventually combined four independent MCMC chains for a total of 4 x 10^9^ iterations after having discarded the first 10% of sub-samples by each chain as burn-in. MCMC convergence and mixing were assessed as described above.

We also used an updated version of the generalised linear model (GLM) extension (29) of the sampling-aware skygrid coalescent model (20) to test for statistical associations between the monthly evolution of the virus effective population size and two climatic covariates: the monthly average of daily mean temperatures and the monthly cumulative precipitation retrieved from the online platform of *Météo France* (meteo.data.gouv.fr). Importantly, these integrated multivariate skygrid-GLM analyses account for uncertainty in effective population size when testing for associations with covariates (60). To test for potential delays in covariate fluctuations being reflected in population dynamics, we also re-conducted all of the skygrid-GLM analyses while specifying a one-month lag period between the monthly covariate values and inferred effective population sizes.

### Inferring the effective reproduction number

We inferred the evolution of the effective reproduction number (R_t_) during the Réunion epidemic (Fig. 1D) using the episodic birth-death-sampling (EBDS) model (21) implemented in the software package BEAST X v1.10.5 (48). This phylodynamic framework models a birth-death-sampling process in which lineages give rise to new infections at a birth rate, become non-infectious at a death rate, and are observed through a sampling rate. Birth and sampling rates are modelled as piecewise-constant across discrete temporal epochs while the death rate is held constant, with lineages removed upon sampling and no intensive sampling events at epoch boundaries. We used a one-year cut-off with 52 grid points (weekly epochs), such that R_t_ is inferred as a step function through time. Priors on the EBDS parameters were specified to be weakly informative while incorporating basic epidemiological constraints. The current birth rate prior was calculated based on an empirical-Bayes approach using the estimated root age and the number of sampled tips, with a log-scale standard deviation set so that the prior spans approximately one order of magnitude above and below the center. The constant death rate prior was informed by an 8-15 day infectious duration window (corresponding to 5-8 days of human infectiousness (56) plus 3-7 days to take into account the mosquito extrinsic incubation period (61)) by specifying a lognormal distribution whose central 95% covers the corresponding bounds. As for the current sampling rate prior, it was set to reflect the present sampling fraction estimated to be around 0.015, mapped to a rate using the calibrated death rate, with moderate uncertainty. Temporal variation in birth and sampling rates was regularised by Gaussian Markov random field priors on the epoch-to-epoch increments of their log rates. For each epoch, the effective reproduction number was computed as the ratio of the birth rate to the sum of the death and sampling rates. For this analysis, we ran and eventually combined five independent MCMC chains for a total of 5 x 10^9^ iterations after discarding the 9.first 10% of sub-samples by each chain as burn-in. MCMC convergence and mixing were assessed as described above.

Similarly to the multivariate skygrid-GLM analyses, we used a GLM extension of the EBDS model to test for statistical associations between the monthly evolution of R_t_ and the same two climatic covariates tested with the skygrid-GLM analyses (i.e. the monthly average of daily mean temperatures and the monthly cumulative precipitation). For compatibility with the skygrid-GLM analyses, the rate grid was set to twelve calendar-aligned monthly epoch spanning the same window. We standardised both climate covariates to zero mean and unit variance across these twelve epochs and parametrized the per-epoch log birth rate as a linear combination of the standardized covariates and an intercept, plus independent Gaussian residuals of fixed variance 0.1. The residual variance was held fixed rather than estimated, since with only twelve epochs an additional smoothing hyperparameter would have been weakly identified by the data. The three regression coefficients were each assigned with a weakly informative Gaussian prior with mean 0 and variance 4, and the Gaussian Markov random field prior on the sampling rate increments was retained from the non-GLM EBDS specification. We placed the GLM on the log birth rate rather than on R_t_ directly because the climate covariates act most naturally on the rate at which new infections are generated, whereas R_t_ is only a derived quantity in the EBDS model. Restricting the covariates to the birth rate, while the death rate stays fixed and the sampling rate carries no covariate dependence, ensures that the GLM coefficient captures the climatic signal in transmission itself, rather than having it absorbed into the sampling or removal processes. And because R_t_ is obtained as the birth rate divided by the sum of the (fixed) death rate and the (smoothed) sampling rate, these coefficients carry over directly to the corresponding associations with R_t_ reported above. To assess whether the climate signal was contemporaneous with or temporally lagged relative to the inferred birth rate, we repeated the analysis with both climatic covariates shifted back by one calendar month, so that each epoch’s log birth rate was regressed on the temperature and precipitation values of the preceding month.

### Phylogeographic reconstruction and analysis

We used the discrete diffusion model (62) implemented in the software package BEAST X v1.10.5 (48) to infer the dispersal history of CHIKV lineages between the island municipalities. As introduced above, this analysis was based on an empirical distribution of trees retrieved from the posterior distribution of a Bayesian time-scaled phylogenetic inference also conducted with BEAST X but without a phylogeographic reconstruction. For this analysis, we ran the MCMC for 2.5 x 10^8^ iterations, sub-sampling every 100,000 iterations, and discarding the first 10% of sub-samples as burn-in. We assessed the MCMC convergence and mixing, as well as retrieved, annotated, and visualised the MCC tree as described above. The resulting phylogeographic reconstruction was visualised using a custom R script adapted from the “seraphim” R package (63, 64) (see the *Code and data availability* section), and we also used the spread.gl program (38) to generate a dynamic visualisation. In order to evaluate the degree of intermixing of the transmission chain across the island, we computed a normalised entropy metric indicating how phylogenetically structured the epidemic was in each municipality. To this end, we computed a previously introduced normalised Shannon entropy measure ranging from 0 (corresponding to a single municipality cluster) to 1 (each sample in the municipality corresponds to a distinct introduction event in that municipality) (37).

In addition, we conducted a discrete phylogeographic analysis using its GLM extension (65) to investigate the contribution of potential predictors to the transition rates of viral lineage between municipalities. With this multivariate GLM analysis, we estimated both the contribution (GLM coefficient) and statistical support (Bayes factor) associated with the different potential predictors; i.e. the geographic distance and professional commuting flow between municipalities, as well as the human population count at the municipality of origin and at the municipality of destination. The geographic distance between two municipalities was computed as the distance, in km, between the centroid points computed from all geo-referenced sampling locations in the respective municipalities. The commuting flow was approximated using 2021 professional mobility data from the French *Institut National de la Statistique et des Études Économiques* (INSEE; www.insee.fr). As for the human population count data, they were computed for each municipality using a 3-arcsecond resolution raster retrieved from the WorldPop database (hub.worldpop.org). Finally, we used an R script adapted from the “spreadStatistic” function of the R package “seraphim” 2.0 (63, 64) to estimate the isolation-by-distance (IBD) signal metric as the Pearson correlation between the patristic (the sum of the branch lengths that link two tip nodes in a phylogenetic tree) and log-transformed geographic distances computed for each pair of tip nodes, and this for each tree sampled from the post-burn-in posterior distribution (39).

## Data Availability

Data Availability: R scripts as well as input and output files related to the different phylodynamic analyses conducted in this study are available at https://github.com/sdellicour/chikungunya_reunion.
Ethical Statement: Samples included in this study consisted of residual serum or plasma from chikungunya cases referred to the NRC for arboviruses in Reunion island. Sequencing data were generated as part of routine surveillance activities within the national public health surveillance program coordinated by the NRC and supervised by the National Public Health Agency (Sante Publique France). As an epidemiological record, consultation with an ethics committee was not required. No additional clinical specimen was collected for the purpose of this study. Human samples were anonymised, with no or minimal risk to patients, in accordance with the European General Data Protection Regulation and the French National Commission on Informatics and Liberty (CNIL).

## Code and data availability

R scripts as well as input and output files related to the different phylodynamic analyses conducted in this study are available at https://github.com/sdellicour/chikungunya_reunion.

## Acknowledgements

We thank Anne-Julie Gourde and Rubens Lhonneur for their assistance with the preparation of the sequencing library, as well as Nicolas M’Nemosyme for his support in the development of bioinformatics pipelines at the NRC Réunion. We also thank *Santé Publique France Réunion* and the *Agence Régionale de Santé – La Réunion* (*ARS La Réunion*) for their support in the management of this epidemic.

## Funding

This work is supported by the ARBOGEN project funded by the MSDAVENIR Foundation (grant agreement n°EM2405-0070381/ANRS-MIE-C24518), and the activity of the National Reference Centers for Arboviruses is supported by the National Public Health Agency (*Santé Publique France*, SPF). KS and SD acknowledge funding from the University of Brussels (ULB, Belgium) internal fund. YS, FM, and MAS acknowledge funding through US National Institutes of Health grants AI153044, AI162611, and AI192139. MSG acknowledges support from the Centers for Disease Control and Prevention, Department of Health and Human Services, under contract NU50CK000626. PL acknowledges support by the Research Foundation *–* Flanders (*Fonds voor Wetenschappelijk Onderzoek* – *Vlaanderen*, FWO, Belgium; grant n°G010326N and G051322N). SD also acknowledges support from the *Fonds National de la Recherche Scientifique* (F.R.S.-FNRS, Belgium; including grant n°F.4515.22), the Research Foundation – Flanders (*Fonds voor Wetenschappelijk Onderzoek – Vlaanderen*, FWO, Belgium; grant n°G098321N), and from the European Union Horizon 2020 project LEAPS (grant agreement n°101094685).

## Author contributions

EF conducted the genomic sequencing; EF, RK, KS, YS, FM, MSG, MAS, PL, and SD analysed the data. YS, FM, MSG, MAS, and PL contributed to the implementation of the analytical pipeline and provided statistical guidance. RK and SD designed the study; RK, XdL, MCJB, and SD supervised the study; EF, RK, KS, and SD wrote the first draft of the manuscript; SD centralised the analytical pipeline and generated the visualisations; all authors edited and approved the final version of the manuscript.

## Competing interests

The authors declare no competing interests.

---

**Figure S1.**
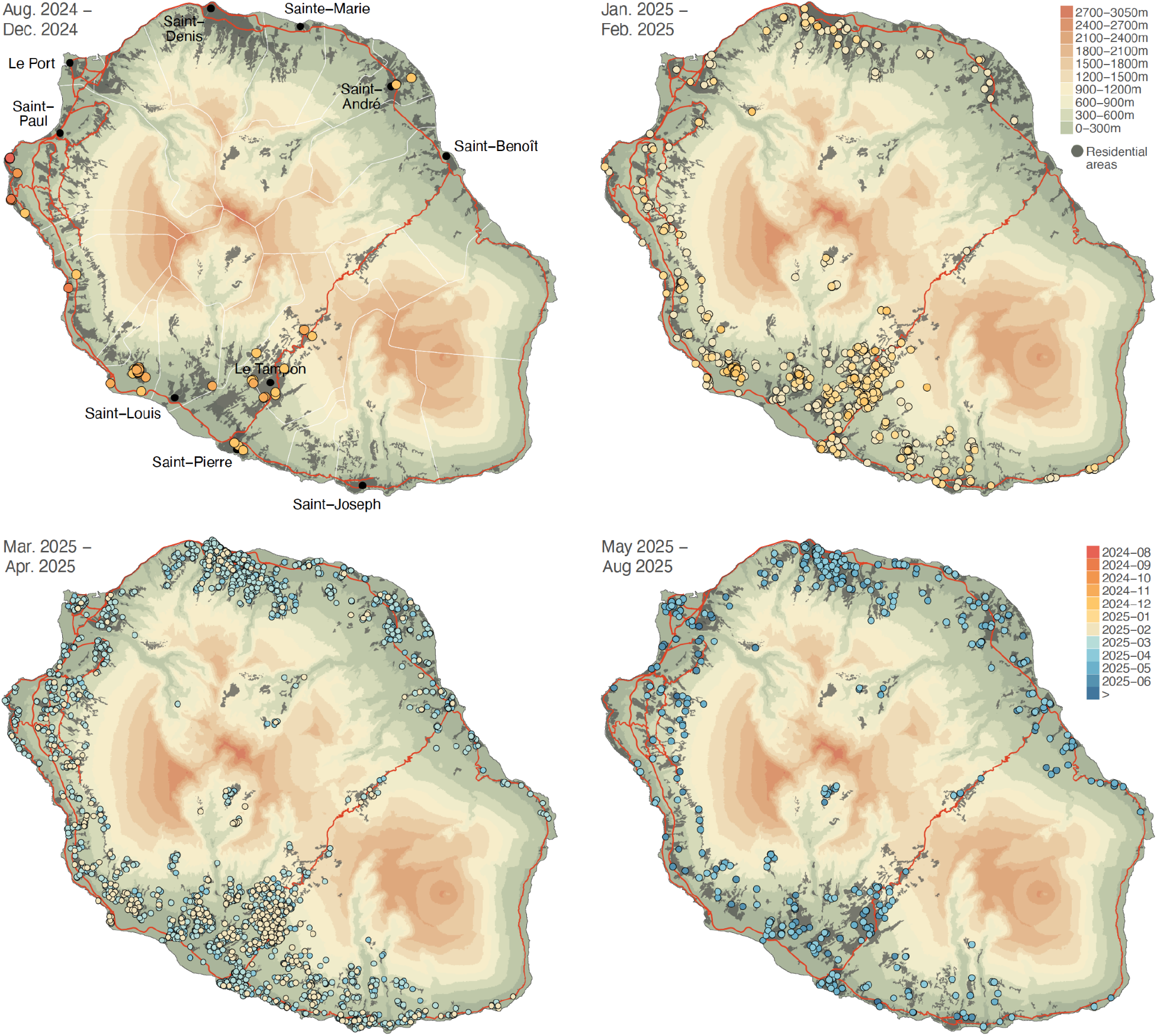
Spatio-temporal distribution of the genomic samples collected and sequenced during the 2024-2025 chikungunya virus (CHIKV) epidemic on Réunion island. The maps correspond to four successive snapshots displaying the distribution of CHIKV genomic samples (coloured dots) collected during the corresponding time periods; each dot being coloured according to the sampling date of the genomic sample. These topographic maps are coloured according to the altitude or in grey when corresponding to residential areas. Red lines correspond to the main roads on the island and, solely reported on the first map, white lines correspond to the borders of the municipalities. On this first map, we also report the position and names of the main urban areas (dark grey dots).

**Figure S2.**
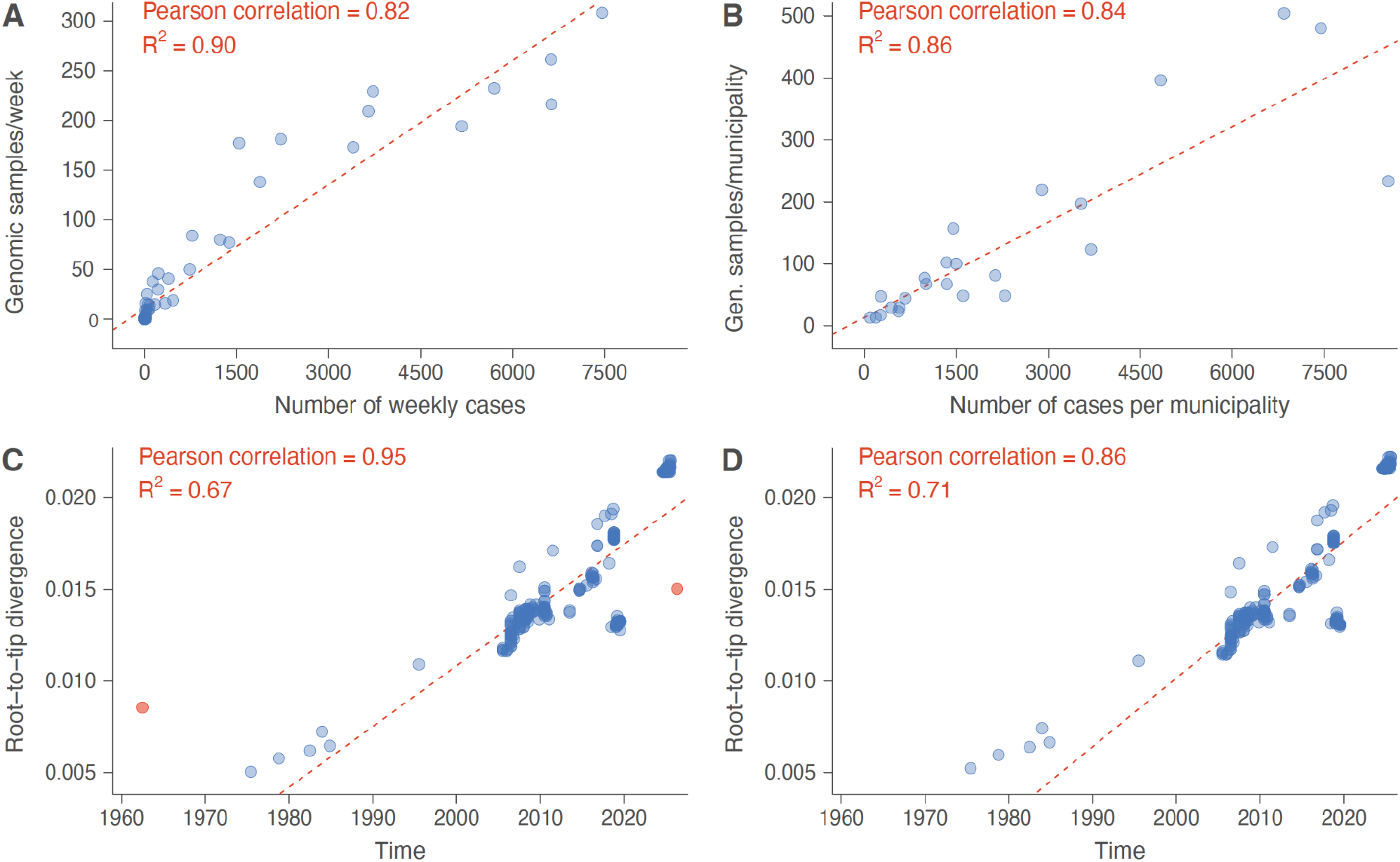
Characteristics of the chikungunya virus (CHIKV) genomic dataset. Linear regression analyses demonstrate the correlation between the number of collected genomic samples and the number of confirmed CHIKV cases per week (**A**), as well as the correlation between the number of collected genomic samples and the number of confirmed CHIKV cases per municipality (**B**). Root-to-tip regression analyses explore the temporal signal associated with the chikungunya virus (CHIKV) genomic alignment, before (**C**) and after (**D**) having excluded two potential outliers (highlighted in red in panel C). “R^2^” refers to the coefficient of determination of each linear regression.

**Figure S3.**
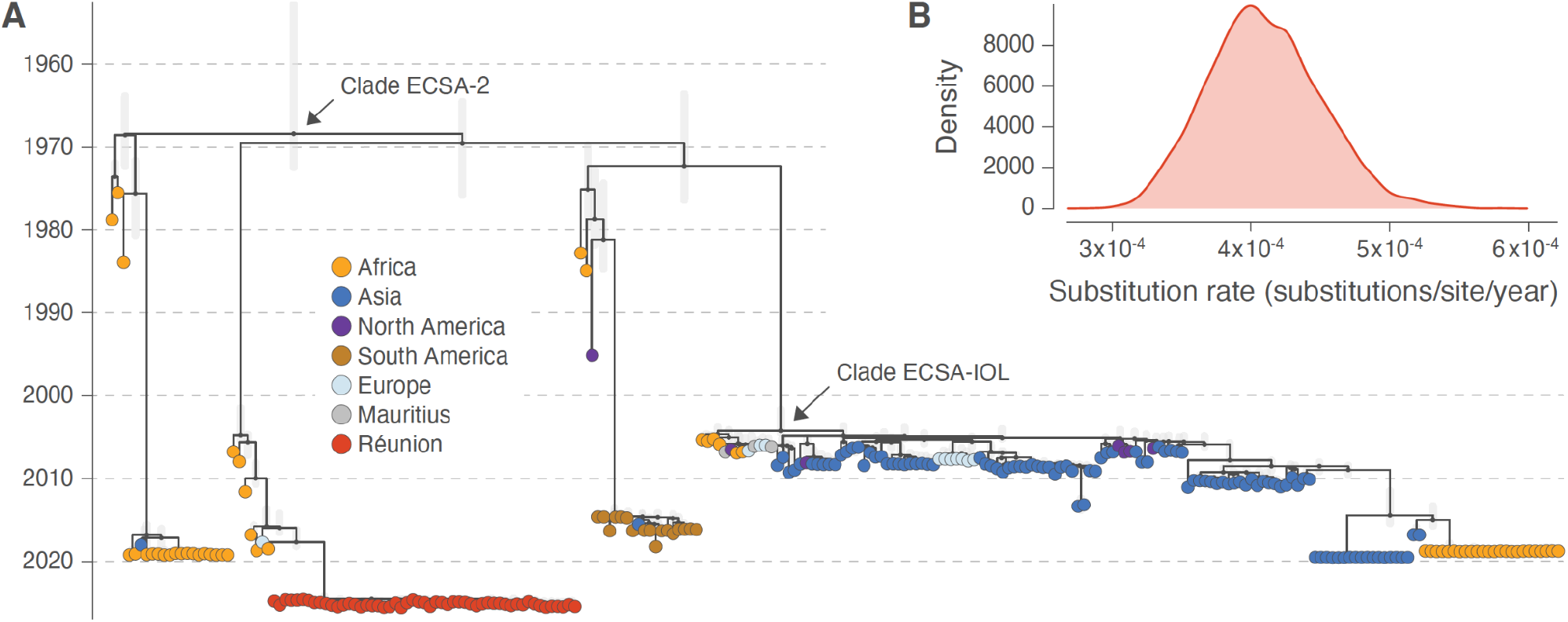
Phylogenetic clustering of the 2024-2025 chikungunya virus (CHIKV) epidemic on Réunion Island. **A**: phylogenetic analysis based on a restricted set of chikungunya virus (CHIKV) genomic sequences collected from Réunion island (*n* = 53, which corresponds to five sequences randomly selected per month) and a set of 198 closely related CHIKV sequences retrieved from GenBank. This phylogenetic analysis was conducted (i) to assess the monophyletic nature of the Réunion clade and thus evaluate the hypothesis of a single introduction event at the basis of the epidemic on Réunion Island, as well as (ii) to estimate a posterior distribution for the substitution rate (**B**). This posterior distribution was used to inform the prior on the substitution rate for all subsequent time-scaled phylodynamic analyses based on the full CHIKV genomic dataset for Réunion island (see, e.g., Figure S4). On the tree, tip nodes are coloured according to their geographic origin and vertical line segments in light grey reflect the 95% highest posterior density (HPD) associated with each internal node age estimate.

**Figure S4.**
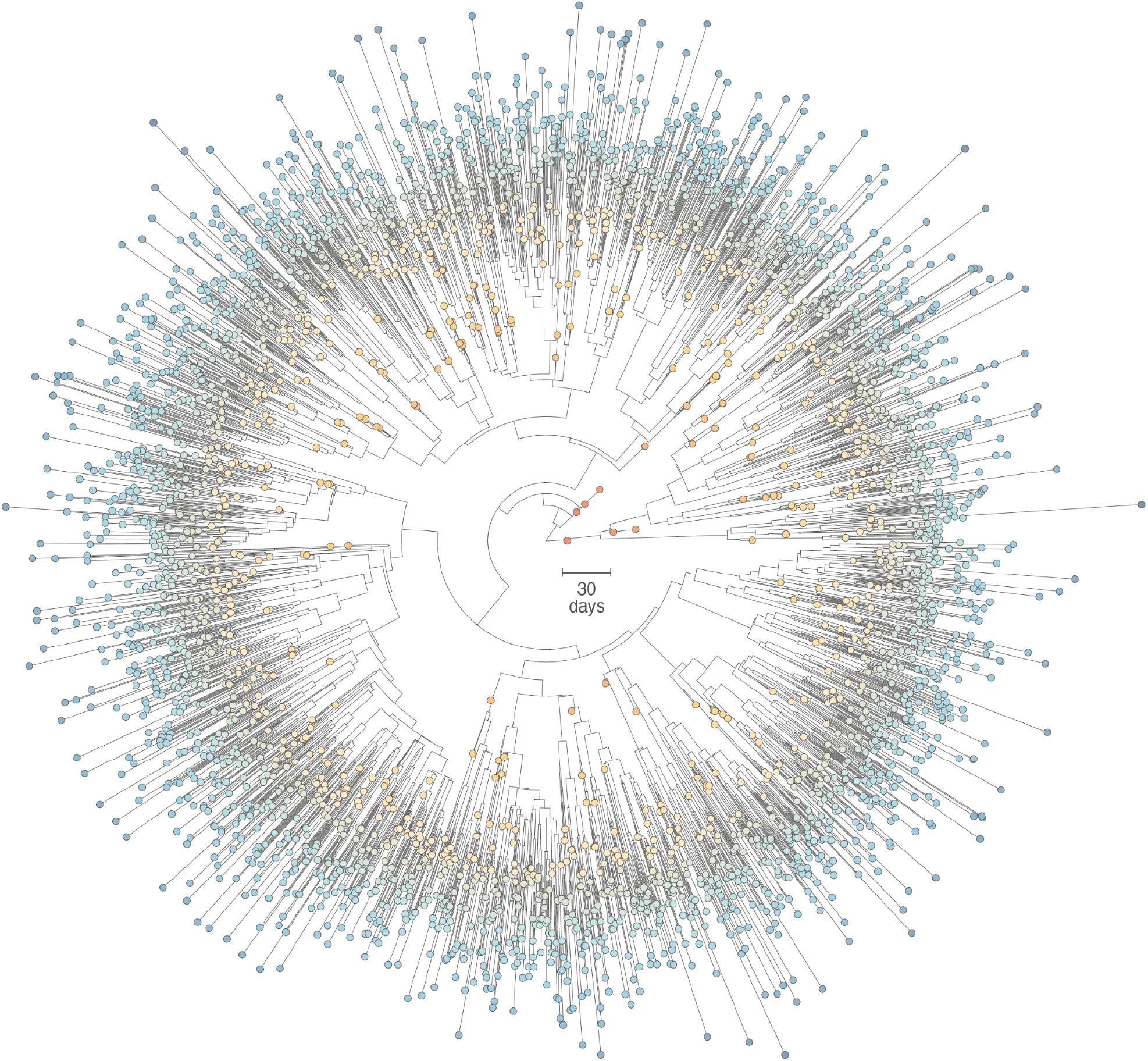
Time-scaled phylogenetic tree inferred for the 2024-2025 chikungunya virus (CHIKV) epidemic on Réunion Island. The reported tree corresponds to the maximum clade credibility (MCC) tree retrieved from the discrete phylogeographic analysis conducted to infer the transition history of viral lineages among municipalities (Fig. 3).

**Figure S5.**
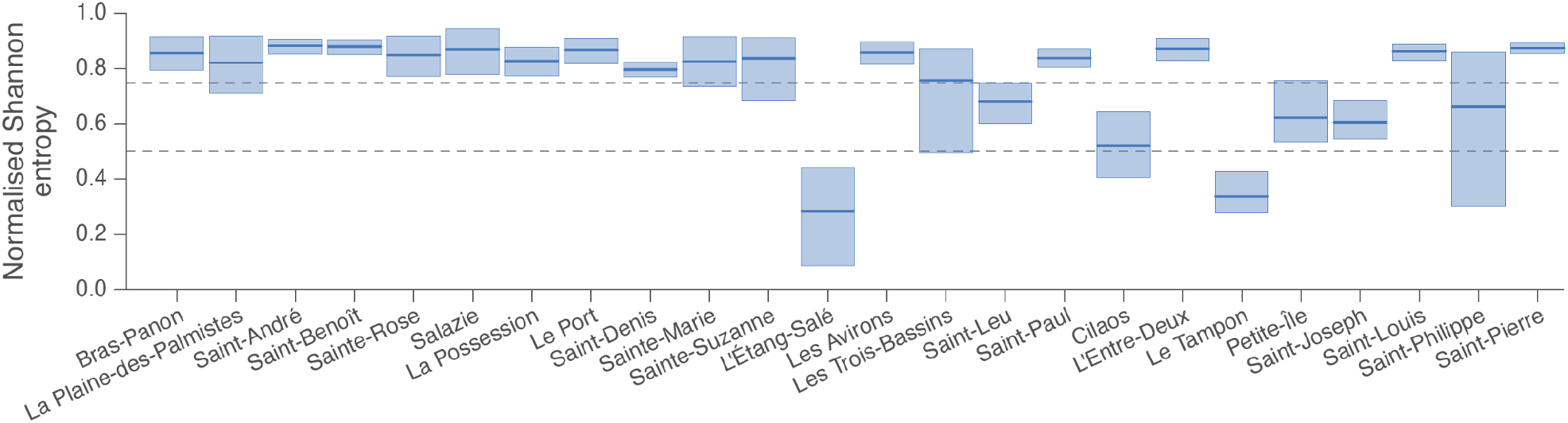
Posterior estimates for the normalised Shannon entropy measuring how phylogenetically structured the epidemic was in each municipality. This metric ranges from 0 (corresponding to a single municipality cluster) to 1 (each sample in the municipality corresponds to a distinct introduction event in that municipality). The thick lines and the boxes correspond to the posterior medians and associated 95% highest posterior density (HPD) intervals, respectively, and the horizontal dashed lines indicate the position of the threshold values of 0.50 and 0.75.

**Table S1.**
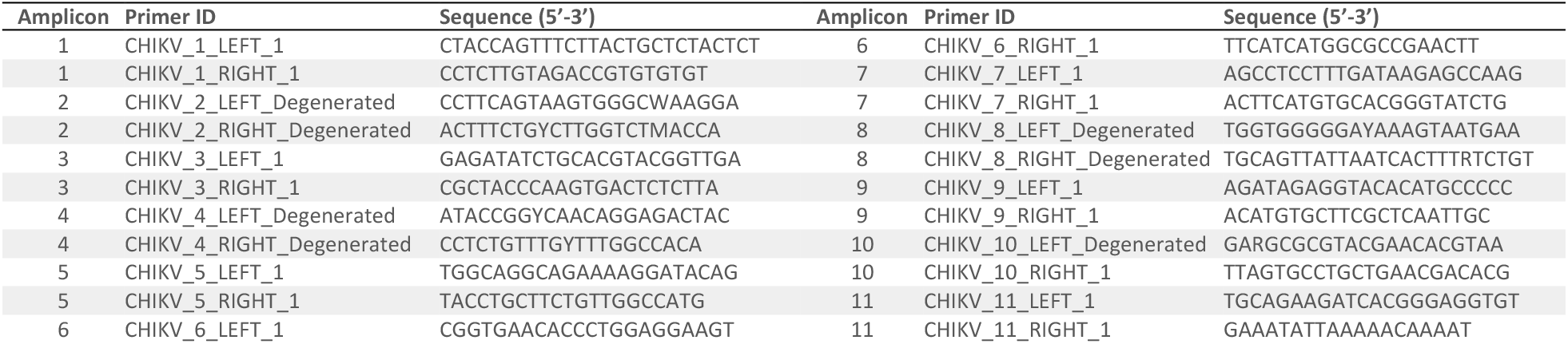
CHIKV primer scheme. Amplicon 2 primers were used at a ×3 concentration to improve coverage and reduce dropout.

## Notes

### Competing Interest Statement

The authors have declared no competing interest.

### Funding Statement

This work is supported by the ARBOGEN project funded by the MSDAVENIR Foundation (grant agreement no. EM2405-0070381/ANRS-MIE-C24518), and the activity of the National Reference Centers for Arboviruses is supported by the National Public Health Agency (Sante Publique France, SPF). KS and SD acknowledge funding from the University of Brussels (ULB, Belgium) internal fund. YS and MAS acknowledge funding through US National Institutes of Health grants AI153044 and AI162611. MSG acknowledges support from the Centers for Disease Control and Prevention, Department of Health and Human Services, under contract NU50CK000626. PL acknowledges support by the Research Foundation - Flanders (Fonds voor Wetenschappelijk Onderzoek - Vlaanderen, FWO, Belgium; grant no. G010326N and G051322N). SD also acknowledges support from the Fonds National de la Recherche Scientifique (F.R.S.-FNRS, Belgium; including grant no. F.4515.22), the Research Foundation - Flanders (Fonds voor Wetenschappelijk Onderzoek - Vlaanderen, FWO, Belgium; grant no. G098321N), and from the European Union Horizon 2020 project LEAPS (grant agreement no. 101094685).

### Author Declarations

This study is based on pathogen genomic data generated within the French national arbovirus surveillance system, which are by now all publicly available on the GenBank database. No additional biological samples were collected for the purpose of this study. The present work represents a secondary analysis of anonymised surveillance data and pathogen genomic sequences and does not fall within the scope of biomedical research involving human participants. In accordance with French law governing public health surveillance activities, formal ethical approval by an institutional review board or ethics committee was not required and was waived. All data processing complied with the European General Data Protection Regulation (GDPR) and the French National Commission on Informatics and Liberty (CNIL).

### Summary of Updates

The main difference with the previous version of our study is that we now also report the results of additional phylodynamic analyses formally investigating the impact of the climatic variables on the dynamic of the epidemics, which confirm statistically supported positive associations with both temperature and precipitation. We have also updated the text to clarify that the population immunity analysis is a theoretical exercise aiming to evaluate whether the population immunity alone could explain the drop of the effective reproduction number (Rt) below one and assess the risk of further virus circulation in human population in upcoming seasons, which does not exclude a potential cumulative impact of other factors, such as climatic factors that are now also formally investigated.

